# Using machine learning-based systems to help predict disengagement from the legal proceedings by women victims of intimate partner violence in Spain

**DOI:** 10.1101/2022.09.29.22280505

**Authors:** Elena Escobar-Linero, María García-Jiménez, María Eva Trigo-Sánchez, María Jesús Cala-Carrillo, José Luis Sevillano, Manuel Domínguez-Morales

## Abstract

Intimate partner violence (IPV) is an actual social issue which poses a challenge in terms of prevention, legal action, and reporting the abuse once it has occurred. In this last case, out of the total of female victims that fill a complaint against their abuser and initiate the legal proceedings, a significant number withdraw from it for different reasons. In this field, it is interesting to detect the victims that disengage from the legal process so that professionals can intervene before it occurs. Previous studies have applied statistical models to use input variables and make a prediction of withdrawal. However, it has not been found in the literature any study that uses machine learning models to predict disengagement from the legal proceedings in IPV cases, which can be a better option to detect these events with a higher precision. Therefore, in this work, a novel application of machine learning techniques to predict the decision of victims of IPV to withdraw from prosecution is studied. For this purpose, three different ML algorithms have been optimized and tested with the original dataset to prove the great performance of ML models against non-linear input data. Once the best models have been obtained, explainable artificial intelligence (xAI) techniques have been applied to search for the most informative input features and reduce the original dataset to the most important variables. Finally, these results have been compared to those obtained in the previous work that used statistical techniques, and the set of most informative parameters has been combined with the variables of the previous study, showing that ML-based models have a better predictive accuracy in all cases and that by adding one new variable to the previous work’ subset, the accuracy to detect withdrawal improves by 7.5%.

## Introduction

Violence suffered by women in intimate partner relationships (IPV) is an important social problem that requires continuous and coordinated intervention from many different spheres. When prevention has not been possible and the violence has already occurred, the complaint can initiate a judicial proceeding. In Spain, only 21.7% of cases in which women have suffered violence from a current partner have been reported, with the woman herself filing the complaint in 80% of cases [1]. Given this low proportion of reported cases, it is important that once the procedure has been initiated, women do not withdraw from it. According to the official data in Spain, withdrawals from legal proceedings in cases of IPV occur between the 9.86% of the complaints, referring to women that abandon prosecution making use of the article 416 of the Criminal Procedure Code [2], and 27.9% of the women that withdraw at any other moment of the legal proceedings [1].

The refusal to continue has important consequences mainly for women, being a predictor of not prosecution [3]. It can have repercussions on the general opinion, including professionals attending these women, increasing the sense of impunity of the accused, but also affecting the credibility of the victims [4]. At the same time, the professionals attending these women may feel that their work has been in vain [5], and the judicial system itself is affected by the increase in its costs. It is therefore vitally important to be able to predict this disengagement, especially if it occurs out of fear or distrust of the justice system, in order to be able to intervene before it happens, advising and supporting women and providing them with the appropriate resources [6].

There have been several studies relating withdrawal of prosecution to different variables (see [5]). Some of the proposed predictive models have used secondary data, mainly drawn from cases dealt with in Domestic Violence Courts (e.g. [3, 7, 8]) and are sometimes limited to cases where only physical violence occurs [9]. More recently, some studies carried out in Spain have overcome these limitations by using data collected directly from women (e.g. [6, 10, 11]). These studies aimed to obtain, through the binary logistic regression (BLR) statistical technique, sufficiently complete and parsimonious models to predict disengagement of legal proceedings, depending on the type of variables used and whether retrospective or prospective data were used. The last of the models developed, as discussed in [12], achieves a correct classification of 83.6% of the cases of disengagements with only six predictors: the victim perceiving that she is not making decisions jointly with her lawyer during the judicial procedure; the woman having thought about the possibility of returning to the abuser after filing the complaint; the protection order previously requested being denied; having contact with the abuser; feeling guilty after filing the complaint; and not receiving psychological support.

With the type of data used in these studies, a low number of experimental subjects, many variables with complex relationships between them, and/or no sufficient a priori knowledge of the relationships between these variables, it is known that Machine Learning (ML) methods could be a better alternative to statistical techniques such as logistic regression [13]. In such cases, an approach based on training classifiers based on a subset of the available data, and then testing this classification using the rest of the dataset, may make better predictions than a classic statistical approach. This is the general aim of this paper: using different ML techniques to study to what extent they can predict women disengagement of legal proceedings, with two specific objectives: a) comparing the predictive power of different ML techniques and with the previously obtained logistic regression models, in terms of general precision, sensitivity, and specificity; and b) comparing the relevant variables detected by the two approaches, ML and BLR, with the possibility to identify new variables that could be useful to predict disengagement using ML methods.

## 1 Background

From a methodological point of view, some studies have already shown the usefulness of using alternatives to regression models (such as neural networks, support vector machines (SVM), or decision trees) given the difficulty, in practice, of complying with the statistical assumptions required for an adequate development and interpretation of the regressions. Likewise, the use of artificial intelligence and, specifically, ML and the procedures indicated above, has interesting advantages over regressions. One of them is the possibility of working even in the presence of multicollinearity among the predictors, or of working with a large group of independent variables or “inputs”, in line with the approach of [14]. This work states that, in the presence of predictors with little predictive power, which are normally eliminated from the models, the accuracy of the prediction can be significantly increased thanks to the aggregation of variables performed by ML. Also, ML techniques could also be more appropriate when it is not possible to assume the existence of multicollinearity between variables [15].

Furthermore, in Social Sciences it seems that the use of ML may be advantageous over regressions in situations in which there are powerful predictors and little “noise” in the data analyzed; even in the opposite case the use of regression is at least as useful as ML both in the construction of the model and in its replication with empirical data [16]. The use of ML brings, in addition, the possibility of model testing by cross validation [17], something that to date has not been done with the regression models proposed by the authors of the papers cited above, whose purpose was to predict withdrawals using regression models. Even though cross validation is possible when using regression models, ML methods ease the quantification of uncertainty as well as making accurate predictions especially with small datasets [17].

Given these benefits, ML procedures have been gaining visibility and strength in Applied Social and Health Sciences. Some works have focused on predicting different types of crimes, and even the location of where they occur, using neural networks and Multilayer Perceptron (MLP) [18]. But there has also been a proliferation of works that aim to predict the type of IPV with Random Forest and SVM classifiers [19] or use neural networks to determine accurate predictors of this type of violence [20]. Furthermore, some ML techniques have been tested to assess the improvement in the accuracy of detection of false positives and false negatives in the assessment of risk for victims of IPV with respect to protocols specifically designed for this purpose [15].

Regarding the decisions made in judicial proceedings, there are also works focused on the use of ML to predict judicial decisions in a wide variety of crimes, such as human rights violations, family and/or civil law cases, and even labor litigation, among others. Rosili’s review [21] lists these and other works that use different Machine Learning techniques to predict judicial decisions for different types of crimes. However, a recent exploratory literature search (see Table 1) has not identified any ML work targeting judicial decisions in the area of intimate partner violence, nor studies using AI for predicting the decisions of victims of this type of violence, including the specific decision to continue or not in a judicial proceeding against the aggressor. Therefore, to the authors’ knowledge, this is the first work using artificial intelligence techniques to predict the decision of victims of gender-based violence not to continue with a legal proceeding initiated against the aggressor.

**Table 1.** Exploratory literature search on Machine Learning to predict withdrawal from prosecution in IPV cases.

|  |  |
| --- | --- |
| Database | Web of Science |
| Date of search | April 8th 2022 |
| Search strategy* | TS=(“intimate partner violence” OR “domestic violence” OR “dating violence” OR “violence against wom*n”) AND TS=(predict* OR forecast*) AND TS=(Withdrawal OR cooperation OR “drop* charge*” OR disengage*) AND TS=(“legal proceedings” OR “criminal justice system” OR “legal cooperation” OR “legal system”) AND TS=(“machine learning” OR “artificial intelligence”) |
| Year of publication** | Any study before 2022 |

*Notes: \*This search found no results;*
*\*\*No other filters were used on Web of Science*

## 2 Materials and Methods

In this section, the components and techniques followed through the development of the study are presented. In the first place, the dataset used is exposed in detail, explaining the data collection process and the summary of participants and variables obtained. In the next sub-section, the complete data pre-processing and processing is presented.

As an introduction, this process involves a first phase of data cleaning, data augmentation and data wrangling, which consist of removing erroneous data, replacing missing values by correct ones following specific techniques, and changing raw data into a more valid format, respectively. In the second phase, a grid search process is carried out for different classifiers, so that the optimization of their hyperparameters can be obtained, and the classification between disengagement and non-disengagement can be more accurate.

The following phase is the study of the most useful variables for each classifier, which is based on understanding the process performed by the AI classifiers to obtain the classification results (as most of them are like ’black boxes’ and the decision process is not clear). To do so, recent works use different techniques that are part of the field known as “Explainable Artificial Intelligence” (xAI) or “Explainable Deep Learning” (xDL). A summary of the most recent techniques can be found in [22].

Finally, the evaluation metrics for each model are obtained to perform a suitable assessment. The graphical abstract of this work can be observed in Figure 1.

**Fig 1.**
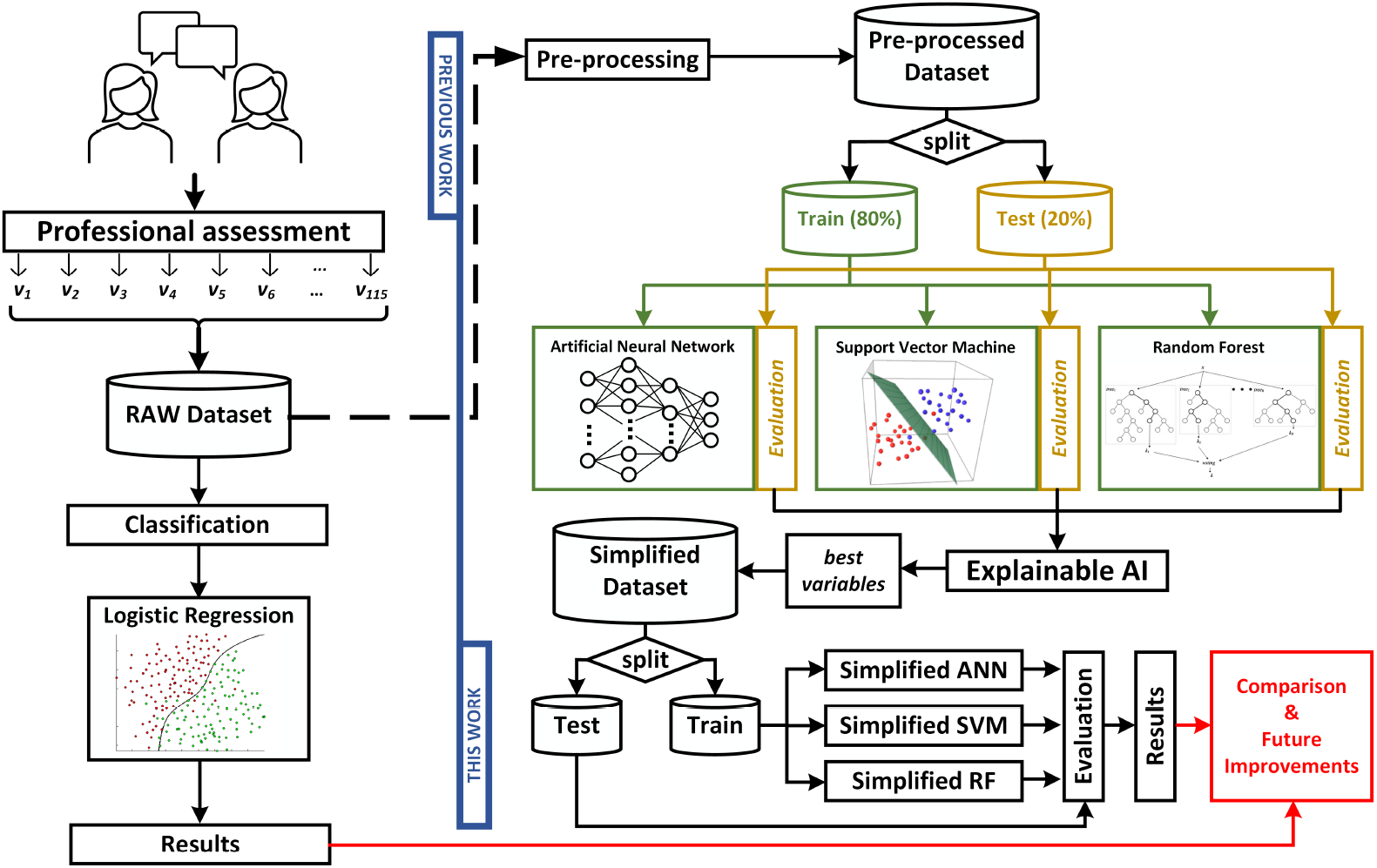
Graphical Abstract. Left side corresponds to the previous work, whose results are compared with the ones obtained in this work (right side).

### 2.1 Dataset

The dataset was obtained from a survey conducted in previous studies (for a more detailed description of the sample and the procedure of data collection [6, 10, 11]). The total sample was 763 women victims of IPV that participated in the legal proceedings against their partners or ex-partners in Southern Spain. They were interviewed in the Andalusian Victims Assistance Service (SAVA in its Spanish acronym), Sheltered Housing, Municipal Centers for Information for Women (CMIM), and some victims help foundations. The average age of the participants was 35.60 (*SD* = 11.09). Out of the total sample, the variable of interest (disengaging or not from the legal proceedings) was first known for the 49.5% of the women whose legal case had been finalized when the data collection started (retrospective data). For the remaining 50.5% of the cases, this variable could not be known until the legal proceedings were ended (prospective data). Overall, 32.2% of women did disengage from the legal proceedings, a higher percentage than in the population because of an intentional attempt to balance the group of participants in both groups.

The variables included in the dataset were 116 (115 after detaching the output variable, named in AI systems as label) before data pre-processing and the posterior creation of dummy variables for analytic purposes (all the variables are detailed on Appendix A). They were extracted from the scientific literature on the topic and interviews with victims and professionals, until information saturation about possible motives to disengage was reached. These variables are grouped into three sets. The first and second sets refer to sociodemographic (e.g., age or number of children) and psychological, emotional and motivational variables (e.g., feelings of guilt after filing the complaint or receiving psychological support), respectively. Both groups were extensively described in [10] and [6]. The third set (see [11]) was related to variables referring to the legal proceedings and professionals involved (e.g., applying for a protection order, whether the protection order was denied).

### 2.2 Pre-processing phase

As mentioned in the introduction of this section, the first phase of the data pre-processing is the data cleaning, data augmentation and data wrangling. This is an essential step for AI classification techniques, since automatic classifiers cannot have any missing values in their input data, as well as the data format has to be appropriate and easy to process to reach a more effective classification. To this end, it is necessary to analyze each sample and apply specific techniques of data pre-processing.

Firstly, the entire dataset has been analyzed, noticing that 32 samples have missing values for the disengagement variable. In this case, as this variable is the actual label to predict and a classifier cannot be trained without labels, the samples have to be removed from the dataset.

Next, it has been observed that each of the 731 resulting samples (763 initials - 32 with missing label) has some missing value in some variables. Moreover, observing the 115 initial variables (the 116th variable was the label), only 20 of them have all the samples, having approximately 80% of variables with any missing value. In view of this, missing values of samples or variables cannot be simply removed, since the final dataset would lose large amount of information and the pre-processing would not be valid in order to obtain a reliable classification. Thus, after removing the samples with missing labels, the variables with more than 15% of missing values were deleted, since it has been shown that higher percentages of missing values could impact severely on the posterior interpretation [23]. After this step, the 115 variables resulted in 47. In order to make the following steps easier, those variables were categorized to numerical values.

At this stage, we enter in the phase of treating missing data. This can be done through data augmentation, with specific methods of value imputation. Specifically, in this study, the mean method was applied, in which missing values are replaced with the average of the variable values [24]. For this step to be more precise, the dataset has been divided into two subsets: one subset containing the samples with disengagement label, and another subset with samples of non-disengagement label. Thus, the average of each variable will be calculated according to the label value, preserving possible information and patterns of the data related to the labels.

The last step of the data pre-processing phase is data wrangling, in which each variable is converted to dummy variables. Dummy variables are the conversion of a categorical variable into a group of variables with values of 0 and 1. In this way, if a categorical variable takes three unique values, there will be three dummy variables to represent it, with values of 1 when the category is present in the corresponding dummy variable. This conversion facilitate the information patterns that the classifier will find in the training process, as well as providing a high flexibility to apply different AI models. In general, dummy variables enable an easy implementation, use and interpretation of the final application [25]. With this step, taking categorical variables to obtain their dummy variables, the final dataset has 93 variables. The summary of the final resulting dataset after data pre-processing is illustrated in Table 2.

**Table 2.** Summary of raw and processed dataset.

|  | Original | Final |
| --- | --- | --- |
| Samples | 763 | 731 |
| Variables | 115 | 93 |

### 2.3 Classifiers

In this work, several classifiers are designed and tested using artificial intelligence techniques to determine whether the decision about abandoning the judicial process in women victims of abuse can be anticipated based on the parameters collected in the dataset.

For this purpose, three known classifiers are used: Random Forest [26], Support Vector Machine [27] and Artificial Neural Network [28]. With all of them, a process of searching for the best possible combination of hyperparameters is carried out (named as “grid search”), performing hundreds of trainings for each classifier and searching for the best classification result on the test subset. The results concerning the best combinations will be shown in the following sections.

Once the best classifiers have been obtained, the most important part of this work is to determine which parameters (variables) in the dataset are the most important to determine the classification. To do this, with the trained classifiers, the percentages of participation of each parameter in the final results are extracted.

For the Random Forest, this can be determined directly by the characteristics of the classifier obtained, but for Support Vector Machine and Artificial Neural Network, this process requires the application of Explainable Deep Learning techniques to elucidate the participation of each parameter in the final result. The xDL technique used is based on the application of the “Anchors” model [29] as a model distillation technique on the dataset. In this technique, commonly applied on images, parts of the image are sequentially removed and their classification behavior is observed.

In our case, the same mechanism will be performed but applied to the parameters of the dataset: the parameter values will be modified one by one by a negative value (unrelated to the information contained) and the changes in the various metrics obtained after classification will be observed. In this way, the parameters that have the greatest impact will be obtained based on those that cause the greatest decrease in the metrics evaluated.

Once the best parameters of each model are known, training will be carried out with various simplified models and combinations between them (including as input only the parameters previously selected). The results obtained after this analysis will allow us to know definitively the importance of these parameters in the final classification.

As a last aspect, and based on the results of a previous study carried out with statistical techniques [6], the parameters selected previously studied will be used with the models developed in this work and combined with the best parameters of this study, in order to extract those parameters that were not taken into account in the previous study but that could improve the final classification (in order to be used in subsequent studies).

The full process is summarized next:

- Phase 1: Obtaining the best classification results using a grid search for each classifier.
- Phase 2: Identifying the best parameters for each classifier applying explainable AI techniques.
- Phase 3: Simplifying the classifiers using only the best parameters as input.
- Phase 4: Using the parameters obtained in the previous works to compare the results.
- Phase 5: Combining the parameters selected in the previous work with the best parameters of this work.

Next, we will begin by describing the various classifiers designed for this work and, subsequently, the techniques and metrics used to evaluate them.

#### 2.3.1 Artificial Neural Network

The first classifier is an Artificial Neural Network, more precisely a multilayer perceptron (MLP). MLP models have an input layer with a number of neurons equal to the number of variables of the dataset, an output layer with a number of neurons equal to the classes to classify and, between these layers, unspecified number of hidden neurons and layers. Moreover, in this MLP architecture, the neurons of a layer are fully connected to the next layer.

The number of hidden layers and neurons is a hyperparameter that has to be chosen. To choose the optimal number, a comprehensive search between different number of neurons and hidden layers is carried out. On the other hand, to reach an accurate prediction, the MLP implements an activation function in each neuron considering the value of the previous connected neuron and a random weight that changes in order to obtain a minimal error in the output neurons. For this computation, several parameters can influence the final value of the actual neuron and with it the final prediction. To obtain an optimal value of the hyperparameters, a grid search process has to be followed, combining different hyperparameters to obtain an optimal model configuration. Firstly, the learning rate is the hyperparameter that determines the update rate of weights in each neuron, known as the step size, and usually takes values between 0 and 1. A smaller learning rate indicates very slow changes on the weights, while higher values indicate the opposite. In the grid search process, learning rate values between 1e-2 and 1e-5 are analyzed. On the other hand, dropout rate is the percentage of neurons randomly ignored by the ML algorithm in the training phase. This is done to prevent the overfitting of the classifier, which is the effect of knowing to well the prior data, with the result that it affects negatively on the predictions over new data. In this way, higher values of dropout rate can result in a more generalized model, although values that are too high can lead to an underfitted model. In the hyperparameter optimization process, the dropout rate varies between 0.1 and 0.3. Once these parameter values that influence on the result of each node are chosen, the ML model can be trained over the dataset. However, it is essential to choose between different batch sizes to obtain a better prediction accuracy. Batch size is the parameter that defines the number of samples that will be introduced in the model before updating any other hyperparameter. Higher batch sizes indicate less changes on the parameters, while with lower sizes more updates are performed, but with the risk of overfitting the model. To choose correctly the batch size of the training step, values between 4 and 32 are analyzed for different parameters combinations.

Overall, different number of hidden layers and neurons with different values of learning rate, dropout rate and batch size are combined in the grid search process to obtain an optimized configuration of all parameters. A total of 1248 combinations are analyzed. Table 3 illustrates the possible values for every hyperparameter.

**Table 3.** Grid Search process for ANN classifier.

|  |  |
| --- | --- |
| Neurons and layers | [2, 3, 4, 5, 6, 7, 8, 9, 10, 11, 12, 13, 2:4, 3:6] |
| Learning Rate | [1e-2, 1e-3, 1e-4, 1e-5] |
| Dropout Rate | [0.1, 0.2, 0.3] |
| Batch Size | [4, 8, 12, 16, 20, 24, 28, 32] |

#### 2.3.2 Support Vector Machine

The second classifier is a Support Vector Machine (SVM), which is an algorithm that searches for a hyperplane in a space of dimension of the number of input variables to separate the data in the different classes. In addition to the hyperplane, it is common to choose the maximal margin hyperplane, which consist of using the closest data points to the hyperplane, known as support vectors, to maximize the distance between those support vectors and the hyperplane [30]. In this way, classification of new data can be more accurate. Another aspect to take into account is that the data is not usually linearly separable and makes difficult to establish a hyperplane separating correctly every data point without having an overfitted model. To overcome this, the model has to add a soft margin that allows the trained model to misclassify some data points. Both margin distance and misclassified points are controlled by C parameter or Regularization parameter. For high values of C parameter, more penalty is applied to each misclassified point, allowing very few misclassifications, and a small distance between the hyperplane and support vectors is chosen to avoid misclassifications. The opposite occurs for smaller values of C parameter. In the grid search process for the optimization of the SVM configuration, C parameter values from 0.01 to 1000 are analyzed.

On the other hand, in SVM the non linearly separable data can be treated with a transformation using kernel functions. An appropriate kernel function maps the original data dimension into a higher dimensional space, known as feature space, so that the data points can be separated more easily. In more detail, the kernel function uses as input the original features and obtains a similarity measure in the new space as output, where similarity refers to the closeness degree of each data point [31].

In this transformation and to control the distance between one point and his neighbour, gamma parameter is used. Higher values of gamma indicates that the closeness of the next point has to be very small so that it can be included in the same class. In the optimization process, gamma parameter values between 1*e −* 4 and 1 are analyzed. Moreover, in this grid search phase, three kernel functions are used: Radial Basis Function kernel (RBF), polynomial kernel and sigmoid kernel.

In short, different values of C parameter, gamma parameter and different kernel functions are combined in 90 possibilities to obtain a SVM classifier with an optimized configuration. Table 4 shows all values used for each hyperparameter.

**Table 4.** Grid Search process for SVM classifier.

|  |  |
| --- | --- |
| **C** | [0.01, 0.1, 1, 10, 100, 1000] |
| **Gamma** | [1, 0.1, 0.01, 0.001, 0.0001] |
| **Kernel** | [RBF, Polynomial, Sigmoid] |

#### 2.3.3 Random Forest

Third and last classifier used is a Random Forest. This model consists of an ensemble of decision trees, where each tree obtains a random vector sampled independently and with the same distribution than every tree in the forest, having uncorrelated tree models [32]. Some settings are essential to obtain an accurate RF model. The first one is the number of estimators, referring to the number of decision trees in the forest. The more estimators the RF model has, the higher accuracy can be reached; however, a high number of decision trees can influence badly on the computing load. In this study, from 100 estimators to 1000 estimators with a step of 100 are applied to the RF model. Another parameter to be considered is the maximum number of features that an individual tree can use. In this case, two size restrictions can be used: the first one, where the tree automatically uses the features that naturally make sense, without having a maximum size; and the second, where the maximum size is the square root of the original features. Higher values of features can affect positively the accuracy, since each tree has more options to consider; however, it can affect negatively on the diversity of each tree.

On the other hand, the maximum depth of each decision tree has to be established, which indicates the splits that the tree have internally. Higher number of splits leads to more information considered, but can also leads to overfitting issues. In the grid search process, the maximum depth analyzed is between 0 and 110 with steps of 10.

The minimum number of samples that can be used to allow the tree to split a node also needs to be considered. With higher number of this parameter, the node has to consider more samples to make a decision and then pass to the next split. In this case, values evaluated in the grid search are from 2 samples to 10 out of the 731 total samples. In a similar way, the number of minimum samples that needs to be on the leaf node has to be established. The leaf nodes are the terminal nodes of the tree, where the decision of belonging to a specific class is done. In this case, the minimum samples between 1 and 4 are analyzed. In both parameters, reaching higher values lead to an underfitting problem, where the algorithm cannot identify patterns on the training data and cannot make any correct classification.

Finally, it is necessary to consider whether or not to apply the bootstrap aggregation or bagging, which is a method that randomly select samples and allow the replacement. Considering all the settings mentioned, the grid search can be carried out over more than 4000 combinations. However, to reduce the computational load, a randomized grid search is realized, with 100 random combinations to find an optimal RF configuration. Table 5 illustrates all hyperparameter values.

**Table 5.** Grid Search process for RF classifier.

|  |  |
| --- | --- |
| **Estimators** | [100, 200, 300, 400, 500, 600, 700, 800, 900, 1000] |
| **Max. Features** | [Auto, Root square] |
| **Max. Depth** | [10, 20, 30, 40, 50, 60, 70, 80, 90, 100, 110, None] |
| **Min. Samples to Split** | [2, 5, 10] |
| **Min. Samples for Leaf** | [1, 2, 4] |
| **Bagging** | [True, False] |

### 2.4 Evaluation procedure

To evaluate the effectiveness in the classification results of a classifier, the most common metrics are used: accuracy (most-used metric), sensitivity (known as recall in other works), specificity, precision, and F1_*score*_ [33]. To this end, the classification results obtained for each class are tagged as ”True Positive” (TP), ”True Negative” (TN), ”False Positive” (FP) or ”False Negative” (FN). According to them, the high-level metrics are presented in the next equations:

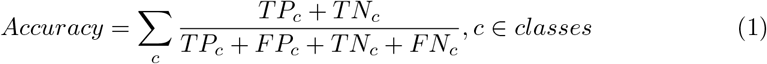

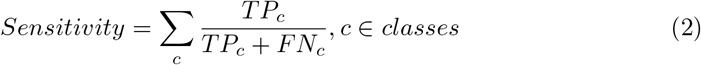

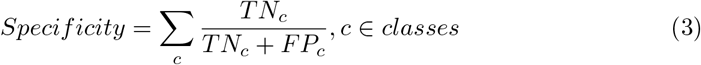

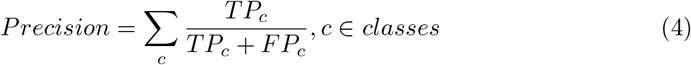

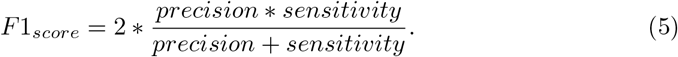

About those metrics:

- Accuracy: all samples classified correctly compared to all samples (see Equation 1)
- Sensitivity (or recall): proportion of values classified as ”true positive” that are correctly classified (see Equation 2)
- Specificity: proportion of values classified as ”true negative” that are correctly classified (see Equation 3)
- Precision: proportion of values classifed as ”true positive” in all cases that have been classified as it (see Equation 4)
- F1_*score*_: It considers two of the main metrics (precision and sensitivity), calculating the harmonic mean of both parameters (see Equation 5)

The above metrics are common to all ML/DL systems. Therefore, the classifier systems developed in this work will be evaluated according to all the metrics detailed in this subsection. Moreover, the results obtained for the classification system, will be compared with the results obtained in previous works.

The results obtained for the previous metrics will be used not only for obtaining the best model for each classifier (phase 1), but to evaluate the classifiers during the xAI application, to compare the different classifiers with the previous work and to obtain the final results of the work (in short, in all of the phases described above, in all of the phases described above).

## 3 Results and Discussion

Following the process detailed phase-by-phase in the previous section, the best results for each classifier are presented below, followed by the application of xAI techniques to extract information from the classifiers and elaborate the final one based on the parameters that provide the most information to the system. Finally, these parameters are combined with those used in the previous work.

### 3.1 Best result for each classifier

After the grid search process, the best candidates can be extracted. Due to the nature of the social problem, the most important aspect is to correctly identify those women who are going to drop out of the judicial process, since these are the cases on which action can be taken to help them. Because of this, the parameter that will mark the goodness of the system will be the sensitivity of the disengagement class.

After the evaluation, for ANN and RF classifiers only one candidate is obtained as the optimized model; however, for SVM classifier, three candidates are obtained with the same results but different parameter configurations.

The best candidate for ANN classifier is formed by one hidden layer with 6 neurons, with a dropout rate of 0.1, a learning rate of 1e-3 and a batch size of 4 samples.

On the other hand, the best RF model consists of 1000 estimators, having in each of them a maximum number of features equal to the square root of original number of features. On the other hand, the minimum samples in each node to allow the split of the tree and the minimum samples in the final nodes are 5 and 2, respectively. Moreover, the bootstrap aggregation method is not applied in this optimized configuration.

Finally, for SVM classifier, three possible optimal configurations are found, named SVM_1_, SVM_2_ and SVM_3_. All of them have in common the application of the RBF kernel, with the gamma parameter value of 0.1. However, SVM_1_ has a C parameter value of 100, SVM_2_ has 1000, and SVM_3_, 10. In Table 6, these optimal model configurations along with the evaluation results are illustrated.

**Table 6.** Best model results for each classifier.

|  | Hyperparameters | Accuracy | Sensitivity | Specificity | Precision | F1-score |
| --- | --- | --- | --- | --- | --- | --- |
| **ANN** | 6 neurons<br>DP: 0.1<br>LR: 1e-3<br>BS: 4 | 91.84 | 0.7576 | 0.9649 | 0.8621 | 0.8065 |
| **SVM<sub>1</sub>** | C: 100<br>Gamma: 0.1<br>Kernel: RBF | 87.76 | 0.8182 | 0.8947 | 0.6923 | 0.75 |
| **SVM<sub>2</sub>** | C: 1000<br>Gamma: 0.1<br>Kernel: RBF | 87.76 | 0.8182 | 0.8947 | 0.6923 | 0.75 |
| **SVM<sub>3</sub>** | C: 10<br>Gamma: 0.1<br>Kernel: RBF | 87.76 | 0.8182 | 0.8947 | 0.6923 | 0.75 |
| **RF** | Estimators: 1000<br>Depth: 60<br>Features: sqrt<br>Split samples: 5<br>Leaf samples: 2<br>Bootstrap: no | 88.44 | 0.7576 | 0.9211 | 0.7353 | 0.7463 |

It can be observed that the best global accuracy value is obtained for ANN model. Moreover, the specificity, precision and f1-score values for ANN classifier is higher than all other models, indicating that the model is better classifying correctly non-disengagement labels, as well as being more consistent and with a better performance over the unbalanced data. However, the model reached a lower sensitivity than SVM models, which is an important metric, since it indicates how well the model classifies the samples as belonging to disengagement class. For this study, it is a better option to have a model that correctly identifies the cases in which the subject disengages from judicial process than it correctly classifies samples as non-disengagement from the process. In this way, all of three SVM models are better options for this prediction, even if the accuracy value obtained is lower than ANN and RF models.

These conclusions can be verified by looking at the confusion matrices of each of these models. The confusion matrix of the best ANN model is shown in Figure 2; the confusion matrix for the three SVM models (the same is obtained for the three of them) is presented in Figure 3; and, finally, the confusion matrix of the best RF model can be observed in Figure 4.

**Fig 2.**
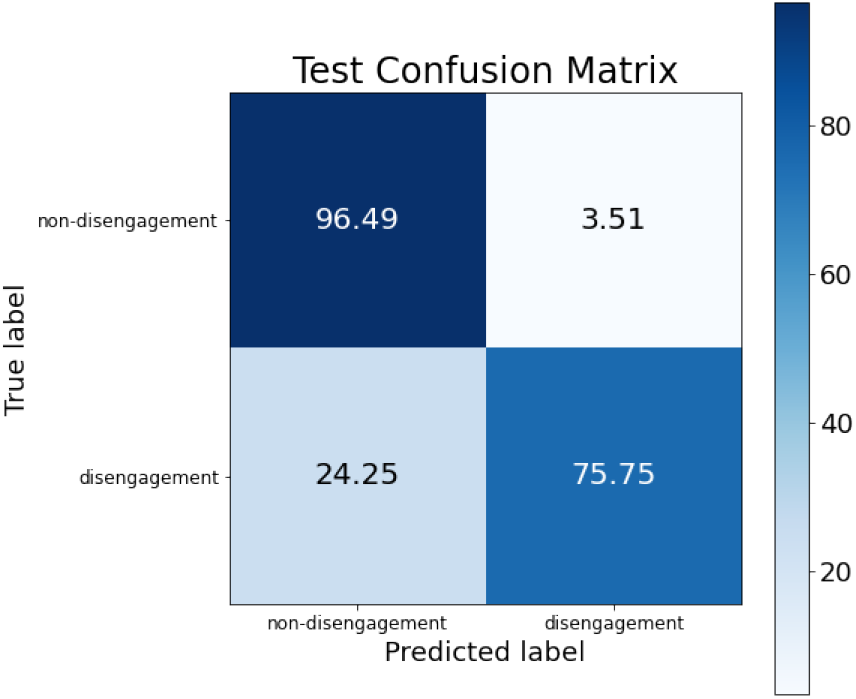
Confusion matrix for the best ANN model.

**Fig 3.**
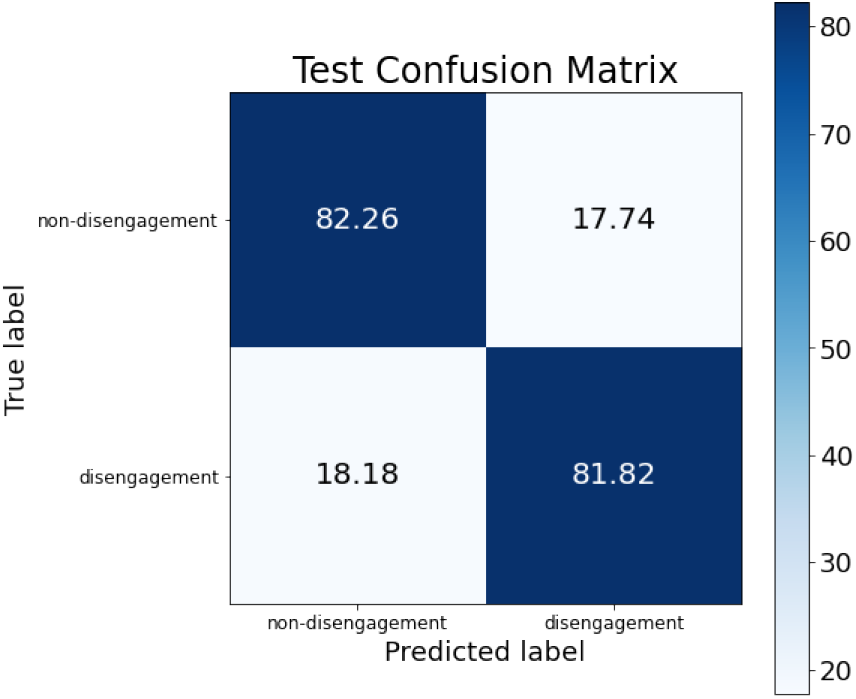
Confusion matrix for the best SVM model.

**Fig 4.**
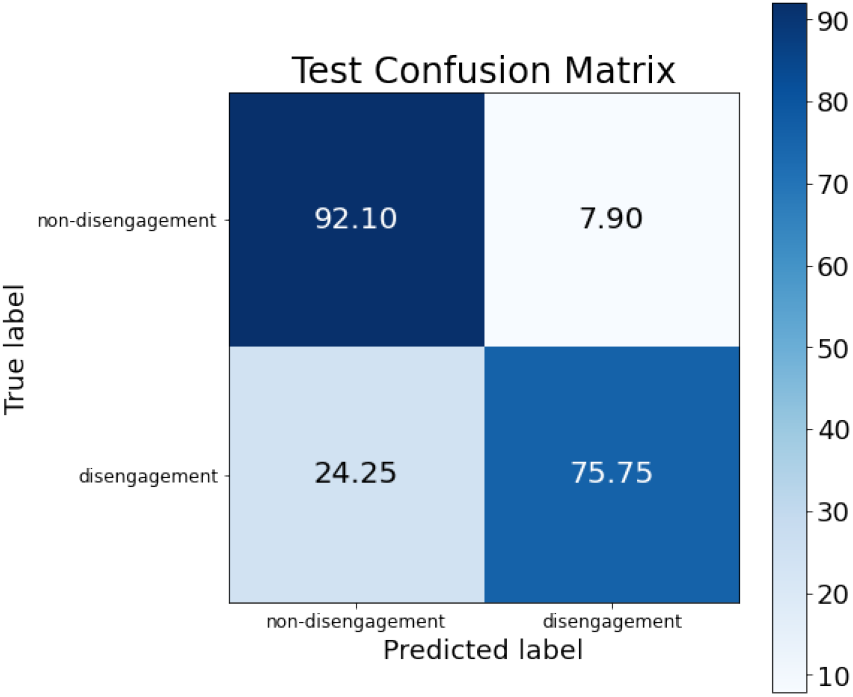
Confusion matrix for the best RF model.

The difference regarding the sensitivity metric detailed before can be observed in the confusion matrices. The percentage of women who drop out of the judicial process and are classified as such is higher in the SVM models than in ANN and RF (see the box at the bottom right of each confusion matrix), obtaining an improvement around 6% (although non-disengagement class obtains an accuracy drop around 14%).

### 3.2 xAI results

Once the best candidates are obtained for each classifier, the most informative parameters are extracted for each model. This can be done through the application of the ‘Anchors’ technique (one of the multiple xAI techniques), as explained in the previous section.

The most important variables for ANN classifier were computed as the difference of accuracy value when the variable is omitted. These results show that 6.4% of the 93 original variables do not have any importance for the classifier (or have a negative influence on it), whereas the 93.6% remaining variables have a minimal importance for the model. In more detail, the majority of variables have an individual influence of changing original accuracy by less than 5%, while a 15% of variables influence in decreasing it by more than 5%. Specifically, 5 of them influence in a decrease of 6.8% of the initial accuracy, and they are considered to be the most informative variables.

These variables are: contact with the aggressor; having a protection order; current questionnaire; considering dropping out of the judicial process and the expectations that the abuser will be imprisoned. However, as explained previously, the variables are initially converted to dummy variables, having one variable of contact with the aggressor and another for not having contact with him. The same occurs for the protection order variable: one variable is obtained for having the order and a second one for not having a protection order. Considering these dummy variables, there are actually 7 variables that influence in the highest decrease of the accuracy.

On the other hand, for the first SVM candidate, the feature importance process showed that 27 variables have a positive influence on the model; while 43 variables do not have any influence on the accuracy result, and the remaining 23 variables influence negatively. Similarly, for both the second and third SVM models, 31 variables have a high importance, 39 do not have any importance, and 23 of them have a negative impact on the model.

Finally, for RF model, results showed that 30 variables have a strong positive impact on the decision of the classifier, while 41 of them does not influenced the model, and 23 of them are even less important, having a negative importance.

These results are summarized in Table 7.

**Table 7.** Variables filtering results obtained for all candidates, classified as positive influence (POS), neutral or minimal influence (NM) and negative influence (NEG).

|  | POS | NM | NEG |
| --- | --- | --- | --- |
| ANN | 7 | 80 | 6 |
| SVM<sub>1</sub> | 27 | 43 | 23 |
| SVM<sub>2</sub> | 31 | 39 | 23 |
| SVM<sub>3</sub> | 31 | 39 | 23 |
| RF | 30 | 41 | 22 |

In the original study, the aim was to determine the minimum number of variables involved in the woman’s decision to abandon the judicial process. If we stick to this premise, we seek the model that requires the fewest variables for this purpose. Due to this circumstance and observing the results in Table 7, the ANN-based model seems to meet this requirement; however, it remains to be determined whether good results are obtained by eliminating all the variables that have not been identified as having a positive influence in each model. This process is carried out as follows.

### 3.3 Simplification of the classifiers

Considering only the most important variables for each classifier (tagged as ‘POS’ in Table 7), a new training phase is carried out by reducing the original dataset to one with the 7 most important variables for ANN model in the first place. With this new dataset, new evaluation metrics were obtained for each of the five best models (see Table 8).

**Table 8.** Evaluation metrics for each classifier with set of ANN’s most important features.

|  | Accuracy | Sensitivity | Specificity | Precision | F1-score |
| --- | --- | --- | --- | --- | --- |
| ANN | 90.48 | 0.8182 | 0.9298 | 0.7714 | 0.7941 |
| SVM<sub>1</sub> | 86.39 | 0.8788 | 0.8596 | 0.6444 | 0.7436 |
| SVM<sub>2</sub> | 87.07 | 0.7879 | 0.8947 | 0.6842 | 0.7324 |
| SVM<sub>3</sub> | 87.07 | 0.7879 | 0.8947 | 0.6842 | 0.7324 |
| RF | 87.76 | 0.7879 | 0.9035 | 0.7027 | 0.7429 |

Following the same steps, others datasets were obtained for the remaining sets of most informative parameters. Table 9 illustrates the results for the classification of each model over the 27 best SVM_1_ variables; Table 10 shows those evaluation metrics obtained with the dataset formed by the 31 most important features for SVM_2_ and SVM_3_ models; and Table 11 presents the results using the dataset composed of the best 30 features obtained with RF classifier.

**Table 9.** Evaluation metrics for each classifier with set of SVM_1_’s most important features.

|  | Accuracy | Sensitivity | Specificity | Precision | F1-score |
| --- | --- | --- | --- | --- | --- |
| ANN | 91.16 | 0.8485 | 0.9298 | 0.7778 | 0.8116 |
| SVM<sub>1</sub> | 89.12 | 0.8182 | 0.9123 | 0.7297 | 0.7714 |
| SVM<sub>2</sub> | 86.39 | 0.8485 | 0.8684 | 0.6512 | 0.7368 |
| SVM<sub>3</sub> | 87.07 | 0.8182 | 0.8860 | 0.6750 | 0.7397 |
| RF | 88.44 | 0.7576 | 0.9211 | 0.7353 | 0.7463 |

**Table 10.** Evaluation metrics for each classifier with set of SVM_2_ and SVM_3_’s most important features.

|  | Accuracy | Sensitivity | Specificity | Precision | F1-score |
| --- | --- | --- | --- | --- | --- |
| **ANN** | 89.80 | 0.7879 | 0.9298 | 0.7647 | 0.7761 |
| **SVM<sub>1</sub>** | 89.12 | 0.8182 | 0.9123 | 0.7297 | 0.7714 |
| **SVM<sub>2</sub>** | 86.39 | 0.8182 | 0.8772 | 0.6585 | 0.7297 |
| **SVM<sub>3</sub>** | 82.99 | 0.7879 | 0.8421 | 0.5909 | 0.6753 |
| **RF** | 89.12 | 0.7273 | 0.9386 | 0.7742 | 0.75 |

**Table 11.** Evaluation metrics for each classifier with set of RF’s most important features.

|  | Accuracy | Sensitivity | Specificity | Precision | F1-score |
| --- | --- | --- | --- | --- | --- |
| **ANN** | 89.80 | 0.7879 | 0.9298 | 0.7647 | 0.7761 |
| **SVM<sub>1</sub>** | 84.35 | 0.6667 | 0.8947 | 0.6471 | 0.6567 |
| **SVM<sub>2</sub>** | 82.99 | 0.6970 | 0.8684 | 0.6053 | 0.6479 |
| **SVM<sub>3</sub>** | 82.99 | 0.7273 | 0.8596 | 0.6 | 0.6575 |
| **RF** | 85.71 | 0.7273 | 0.8947 | 0.6667 | 0.6957 |

As can be seen in the previous tables, the best classification results are obtained by training the new reduced ANN model using as input only the most important variables of the previous SVM_1_ model (27 parameters), presented in the first line of Table 9.

The overall results show that the resulting accuracy is practically identical than the best obtained in the initial models, but a very interesting phenomenon occurs: by reducing the parameters that influenced negatively or minimally the initial model, the sensitivity value for the new ANN model increases drastically and outperforms the initial SVM models. Thus, the metric used to measure the goodness-of-fit of the system improves by almost 10% with respect to the initial ANN model.

However, the number of entries is a negative aspect to take into account, as 27 parameters is a very large number for the initial social study. But, if we look at the results obtained by the new ANN model trained with the best parameters of the initial ANN model (only 7 parameters) shown in the first line of Table 8, the results do not differ much from those of the best case (a reduction of less than 0.7% in accuracy and 3% in sensitivity). Although this is an aspect to be taken into account, when looking for the simplest possible model, this model can be a strong candidate.

So, after this simplification process, the ANN model is the best candidate in both cases (with 7 and 27 parameters). Their confusion matrices can be seen in Figure 5 for the 7-parameter model, and in Figure 6 for the 27-parameter model.

**Fig 5.**
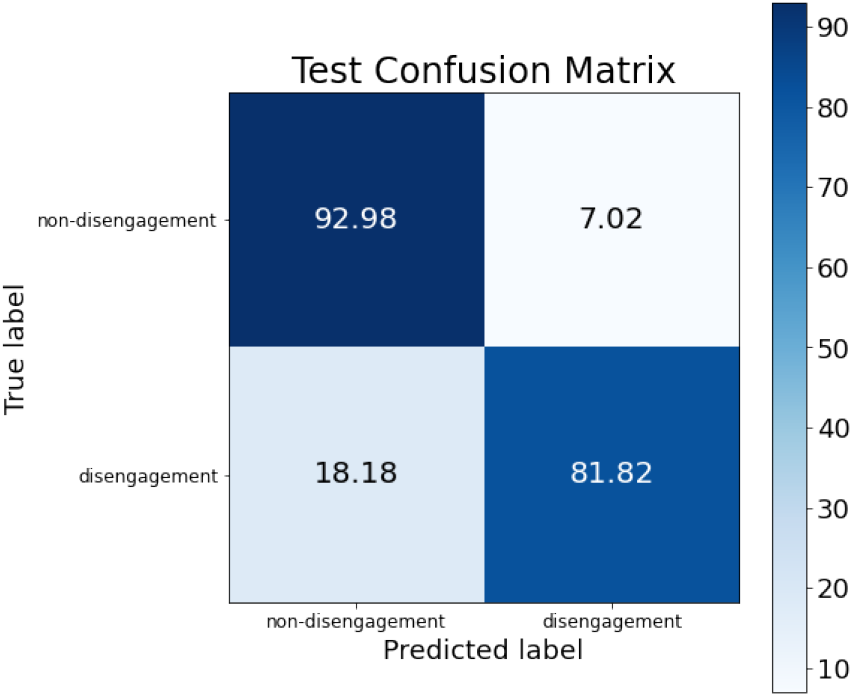
Confusion matrix for ANN model trained with the best 7 parameters from the initial ANN model.

**Fig 6.**
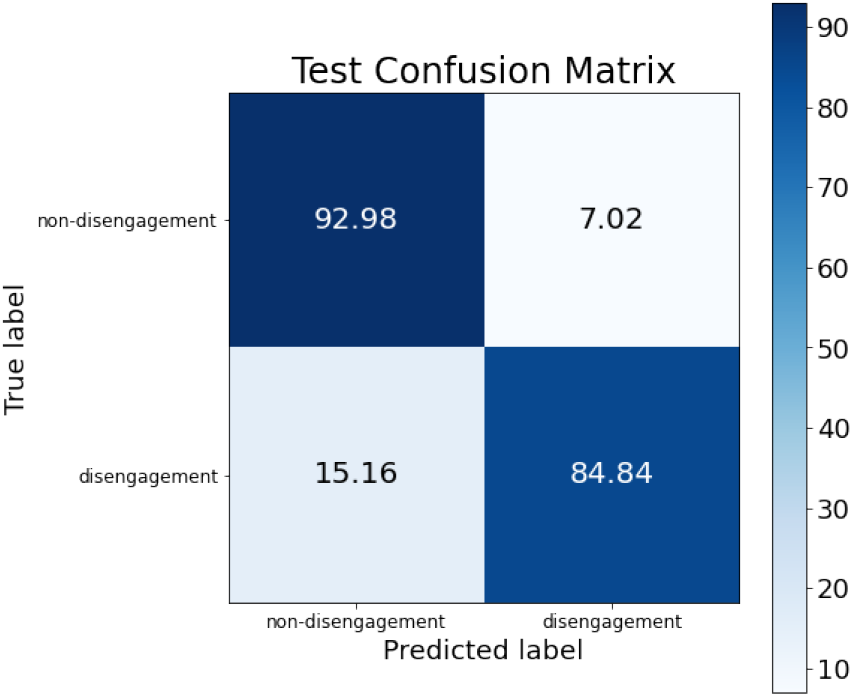
Confusion matrix for ANN model trained with the best 27 parameters from the initial SVM_1_ model.

If we look at the main differences between the two simplified models, it can be appreciated that the percentage of samples correctly classified for the non-disengagement class is exactly the same, but for the disengagement class there are differences: the 27-parameter model correctly classifies 84.84% of the dataset’s samples for this class, while the 7-parameter model classifies 81.82% (this represents a difference of 3% between them).

However, as the dataset used has a not very large number of samples and the test subset represents a 20% of the total number of samples, the normalized difference of 3% obtained before is only an absolute difference of one sample, i.e. the 27-parameter model correctly classifies one more sample than the 7-parameter model.

This difference in classification is not as significant as the difference in classifier complexity: to correctly classify one more sample requires a classifier with 4 times the number of entries. Thus, the most promising candidate in terms of accuracy and complexity is the ANN-based classifier with 7 input parameters.

### 3.4 Classifiers with best parameters from the previous work

After evaluating all best classifiers with the most important variables, next step is to evaluate those ML models with the set of most significant variables obtained in the previous work of [34], in order to compare with the same technology both sets of parameters. The final parameters selected in that work were: receiving psychological support; contact with the abuser; thinking of going back with the aggressor; feeling guilty; having a protection order and perceiving that the decisions of the judicial procedure are not made jointly with her lawyer.

Results of evaluation metrics obtained for each model for this set of parameters is shown in Table 12.

**Table 12.** Evaluation metrics for each classifier with set of previous work’s most important features.

|  | Accuracy | Sensitivity | Specificity | Precision | F1-score |
| --- | --- | --- | --- | --- | --- |
| **ANN** | 87.07 | 0.6667 | 0.9298 | 0.7333 | 0.6984 |
| **SVM<sub>1</sub>** | 86.39 | 0.6970 | 0.9123 | 0.6970 | 0.6970 |
| **SVM<sub>2</sub>** | 86.39 | 0.6970 | 0.9123 | 0.6970 | 0.6970 |
| **SVM<sub>3</sub>** | 87.07 | 0.7273 | 0.9123 | 0.7059 | 0.7164 |
| **RF** | 86.39 | 0.7273 | 0.9035 | 0.6857 | 0.7059 |

As can be observed the accuracy value obtained is lower than the one achieved before, and the sensitivity metric has fallen sharply to values of around 66-72%. For the best case of all of them (SVM_3_), the confusion matrix obtained is shown in Figure 7.

**Fig 7.**
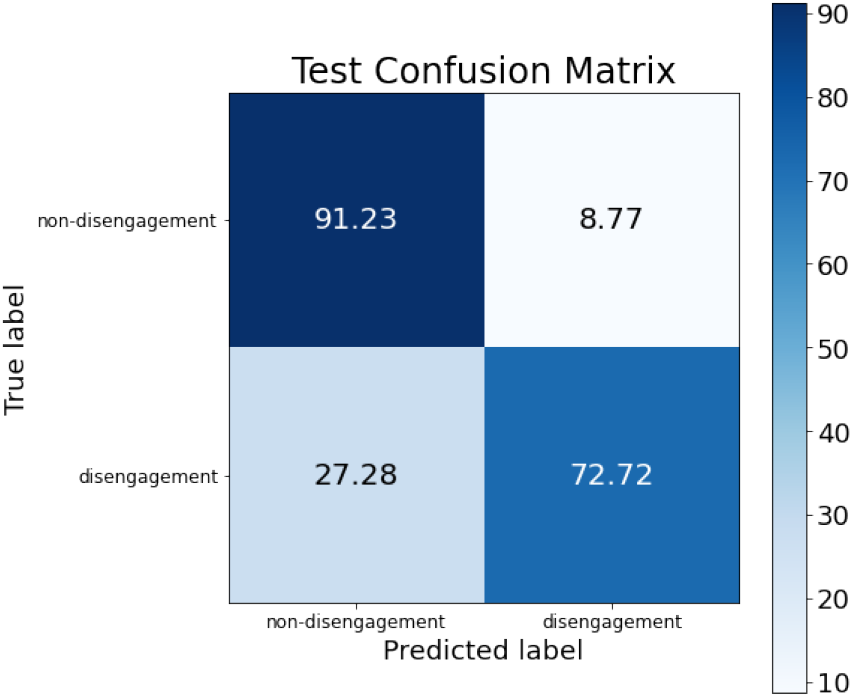
Confusion matrix for SVM_3_ model trained with the parameters used in the previous work.

Compared with the previous simplified model, the non-disengagement class accuracy drops only 1.75%, but the main problem remains in the disengagement class, whose accuracy value drops more than 12%.

These results indicate that in the previous work certain parameters were discarded from the dataset, and those parameters may improve the classification rate of the system. With this information, in the last subsection the difference in the parameter selection between the previous study and this new study is studied, and some combinations between those parameters are performed in order to determine which parameters (initially discarded in the previous work) can improve the classification.

### 3.5 Parameters combination

The last step of this comprehensive analysis of variables includes the combination of the most important variables obtained for the different classifiers, together with those variables with a significant value obtained in the previous work.

If we observe the similarities between the variables selected in the previous work and the most significant variables obtained in the “simplification of classifiers” subsection of this study, we obtain the results shown in Table 13.

**Table 13.**
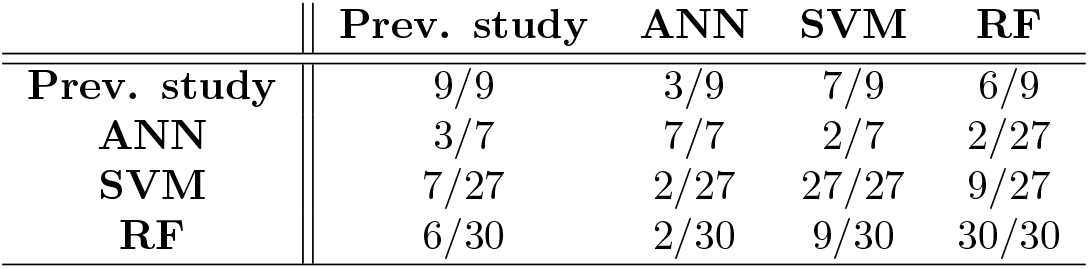
Variable comparison between previous study and the classifiers optimized for this work. Information presented in fraction mode, where the denominator represents the number of variables used for the row’s system, and the numerator represents the number of variables of the row’s system that matches the column’s system

The first column of Table 13 indicates the number of variables from each system that matches the variables selected in the previous work. For the best candidate selected in this work (ANN), there is a 43% of coincidence in the variables selected.

The next study combines the variables used in the previous works with the new variables used in this study for each candidate. If the new systems were trained adding the variables one by one, millions of combinations would have to be performed; so the combinations are done by grouping the new variables in four subsets:

- Subset 1: it contains those variables common in the three new systems (ANN, SVM and RF) that are not used in the previous work.
- Subset 2: it contains those variables only used in ANN that are not used in the previous work.
- Subset 3: it contains those variables only used in SVM that are not used in the previous work.
- Subset 4: it contains those variables only used in RF that are not used in the previous work.

So, four new classifiers are trained combining the variables used in the previous work with the ones of each subset (named as “combined classifier” 1-4, or CC1-4 to simplify). The results obtained for testing those combinations in the ANN model are presented in Table 14.

**Table 14.** Combined classifiers’ results. Each row shows the results of the classifier trained with the previous work’s variables and the ones that contains the subset indicated in that row.

|  | Accuracy | Sensitivity | Specificity | Precision | F1-score |
| --- | --- | --- | --- | --- | --- |
| CC1 | 90.48 | 0.8182 | 0.9464 | 0.7714 | 0.7941 |
| CC2 | 87.76 | 0.7272 | 0.9210 | 0.7272 | 0.7272 |
| CC3 | 90.48 | 0.8182 | 0.9464 | 0.7714 | 0.7941 |
| CC4 | 89.12 | 0.7575 | 0.9298 | 0.7353 | 0.7462 |

As can be observed in Table 14, the systems CC1 y CC3 obtain the same results. CC1 corresponds to the combination of the previous work’s variables and the common variables of the three new systems; while CC3 corresponds to the combination of the previous work’s variables and the variables only used for the SVM system.

The main difference between both of them is that CC1 includes only 2 new variables, while CC3 includes 21 new variables. Looking for the less complex system, the best candidate in this case is CC1.

The two new variables used in CC1 are: “plans to abandon” and “current questionnaire”. As they are only two, the next step is to analyse the inclusion of these variables one by one in order to know if both variables improves the previous work results or, in other case, only one of them does.

Table 15 illustrates the results of the classification combining the variables used in the previous work with the common variable “plans to abandon”.

**Table 15.**
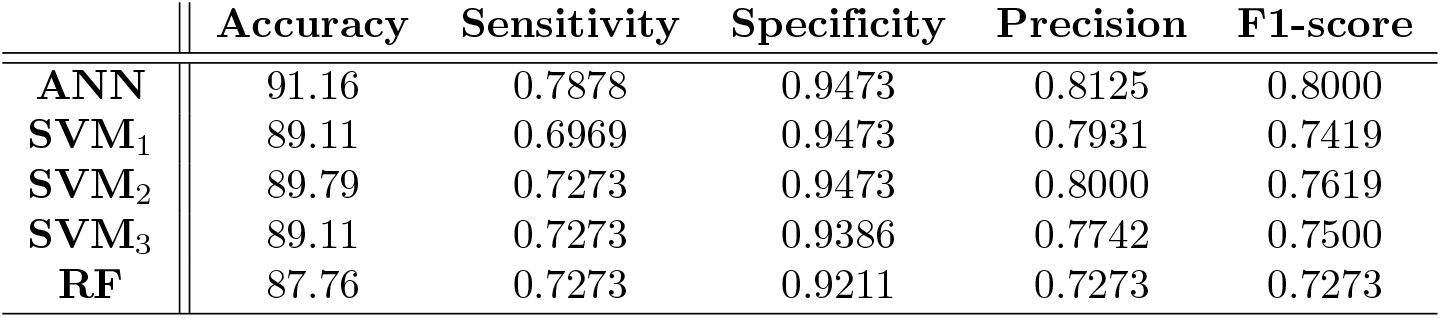
Evaluation metrics for each classifier with set of previous work’s most important parameters combined with “plans to abandon”.

|  | Accuracy | Sensitivity | Specificity | Precision | F1-score |
| --- | --- | --- | --- | --- | --- |
| ANN | 91.16 | 0.7878 | 0.9473 | 0.8125 | 0.8000 |
| SVM<sub>1</sub> | 89.11 | 0.6969 | 0.9473 | 0.7931 | 0.7419 |
| SVM<sub>2</sub> | 89.79 | 0.7273 | 0.9473 | 0.8000 | 0.7619 |
| SVM<sub>3</sub> | 89.11 | 0.7273 | 0.9386 | 0.7742 | 0.7500 |
| RF | 87.76 | 0.7273 | 0.9211 | 0.7273 | 0.7273 |

Table 16 illustrates the results of the classification combining the variables used in the previous work with the common variable “current questionnaire”.

**Table 16.**
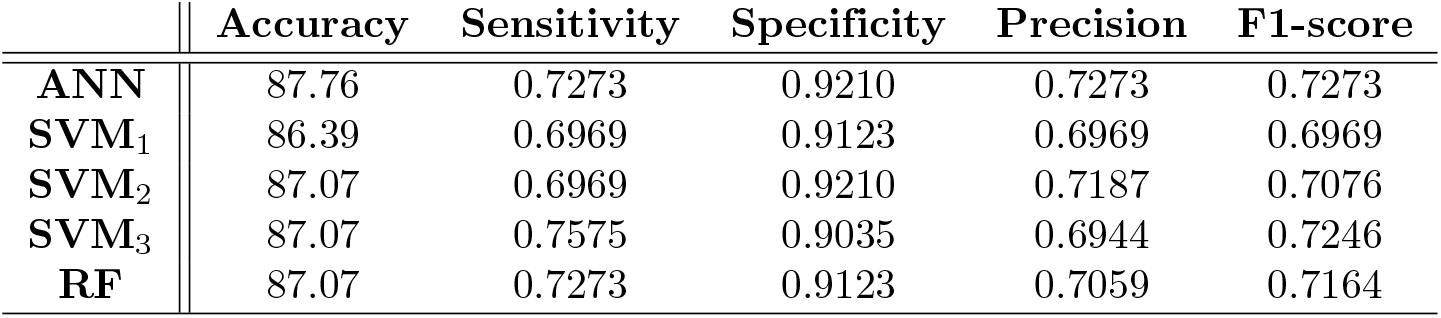
Evaluation metrics for each classifier with set of previous work’s most important parameters combined with “current questionnaire”.

|  | Accuracy | Sensitivity | Specificity | Precision | F1-score |
| --- | --- | --- | --- | --- | --- |
| ANN | 87.76 | 0.7273 | 0.9210 | 0.7273 | 0.7273 |
| SVM<sub>1</sub> | 86.39 | 0.6969 | 0.9123 | 0.6969 | 0.6969 |
| SVM<sub>2</sub> | 87.07 | 0.6969 | 0.9210 | 0.7187 | 0.7076 |
| SVM<sub>3</sub> | 87.07 | 0.7575 | 0.9035 | 0.6944 | 0.7246 |
| RF | 87.07 | 0.7273 | 0.9123 | 0.7059 | 0.7164 |

In the previous tables, three main findings are presented:

- Point 1: the inclusion of the variable “plans to abandon” improves the previous work classification from 87.07% to 91.16% (more than a 4%), obtaining the same global accuracy that the one presented in Table 9 for the best ANN parameters.
- Point 2: the inclusion of the variable “current questionnaire” does not provoke a significant improvement (only a 0.69%).
- Point 3: the inclusion of both variables improve the previous work classification results, but obtains worse results that the ones obtained by including only the variable “plans to abandon”.

So, after analysing the consequences of including the new two variables, the best solution is obtained by combining the previous work variables with the variable “plans to abandon”. The final results are detailed in its confusion matrix (see Figure 8).

**Fig 8.**
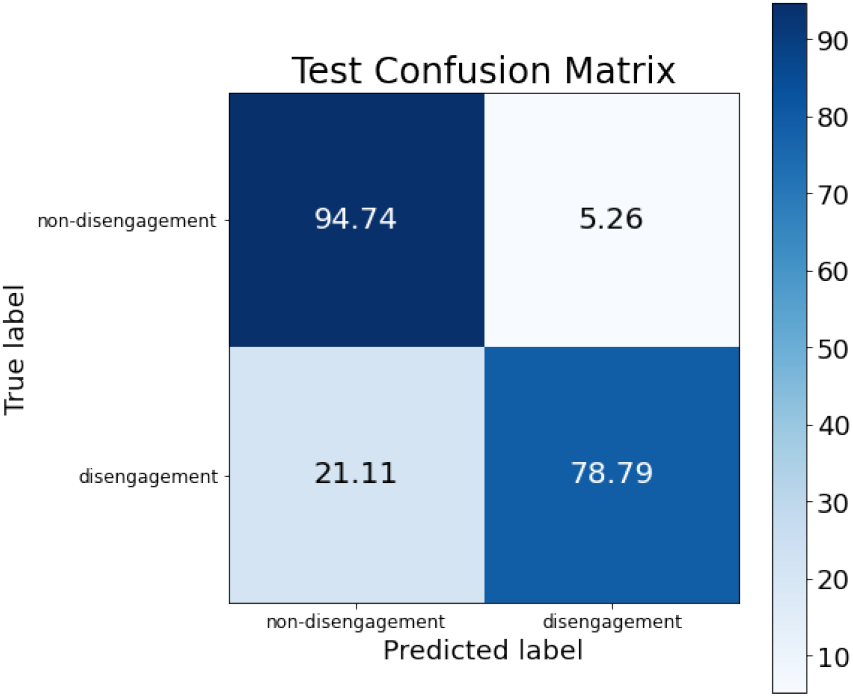
Confusion matrix for the final model obtained by the combination of the previous work variables and the new variable “plans to abandon”.

## 4 Conclusions

In this work, different Machine Learning models are applied to predict victims of gender-based violence disengagement from the legal proceedings, with the general purpose of obtaining a reliable support system so that the professionals can intervene 577 before the withdrawal occurs.

For this purpose, three classifiers with different sets of the original dataset are tested, optimizing previously their hyperparameters and applying explainable AI techniques to obtain a subset of the most informative variables. This phase aims to identify the model that best classifies disengagement by the victim by using the minimum and most informative set of input parameters. Results have shown that ANN-based classifier is the best candidate identifying 7 variables as the most informative and obtaining an accuracy of 91.16%.

Moreover, this work intents to apply the ML classifiers to the set of variables used in the previous study (where a logistic regression model was used), so that it can be demonstrated that AI models obtains better accuracy results that the ones obtained in the previous study. The best result obtained in this case was for SVM-based classifier with an accuracy of 87.07%, which is an improvement of more than 3%, compared to the accuracy of 83.6% obtained in the previous work.

In the last phase, by applying the different classifiers to the combination of the set of most informative variables with those obtained in the previous work, results showed that by adding one new variable (“plans to withdraw”) to the previous work’ subset,the accuracy improves by 7.5%. Adding this variable is particularly useful since it allows professionals to intervene if a woman discloses such intention.

Overall, results obtained in this work showed that using ML-based classifiers to this problem and, thanks to the comparison work and the xAI studies, using one more variable to the previous work’s set, the accuracy results are improved substantially. Thus, in future studies, this new variable will be taken into account to reach more reliable predictions. However, it is worth noting that the dataset used for the detection of disengagement of the judicial process consists of 731 victims, which remains in a low number of samples for the model to make consistent and reliable predictions with new data. Therefore, future works will focus on extending the original dataset and make predictions by adding the new variable to the input parameters. Despite this limitation, the use of ML and the addition of new variables to the original works provides an additional benefit regarding the conceptualization of IPV victims. Such previous works argued the principle of parsimony which, despite its usefulness, it may entail the risk of reducing victims that withdraw from prosecution to specific features and the wrong idea of an existing IPV victim profile. The different ML models implemented here have enriched the characteristics of the phenomenon studied and eased the understanding of the complexity of IPV and victims’ decision-making processes.

Finally, future applications sought with the application of ML models are the possibility of obtaining systems that allow more accurate detection of whether the victim will end up abandoning the judicial process. Thus, through these support systems for professionals, it will be possible to detect some of the characteristics of women who are likely to withdraw prosecution and therefore provide them with more professional help to prevent them from doing it. This could be done by developing a mobile or web application in which the predictor model is integrated and, after including the answers to the initial forms answered by the victims before starting the judicial process, the professionals could obtain a prediction about the possible withdrawal and recommendations to for action. In any case, and in line with [15], any professional tool based on artificial intelligence that is implemented must necessarily be complemented with adequate professional assistance, especially when working with victims of a crime such as IPV.

## Data Availability

Data cannot be shared publicly in this work because of being collected and shared in a previous work.

## Acknowledgments

This work has been partially supported by ”Fondo Europeo de Desarrollo Regional” (FEDER), ”Consejería de Igualdad y Bienestar Social” (1071/0453) and ”Consejería de Economía, Conocimiento, Empresas y Universidad” (POSDOC-21-00604) of the Junta de Andalucía, and under Programa Operativo FEDER 2014-2020 (Project US-1264911) as well as by the Telefonica Chair ”Intelligence in Networks” of the Universidad de Sevilla.

## A Variables in the dataset before the data pre-processing

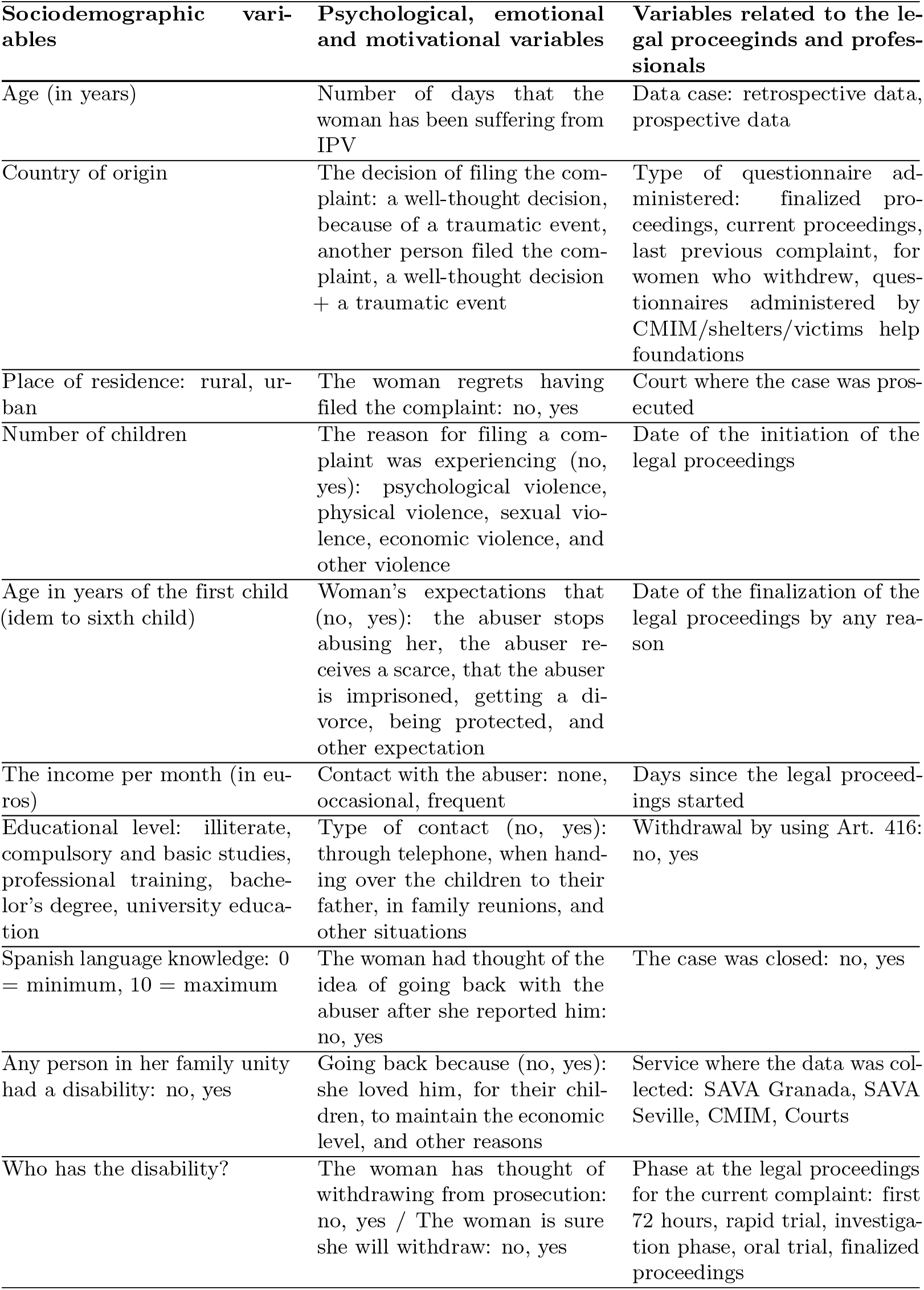
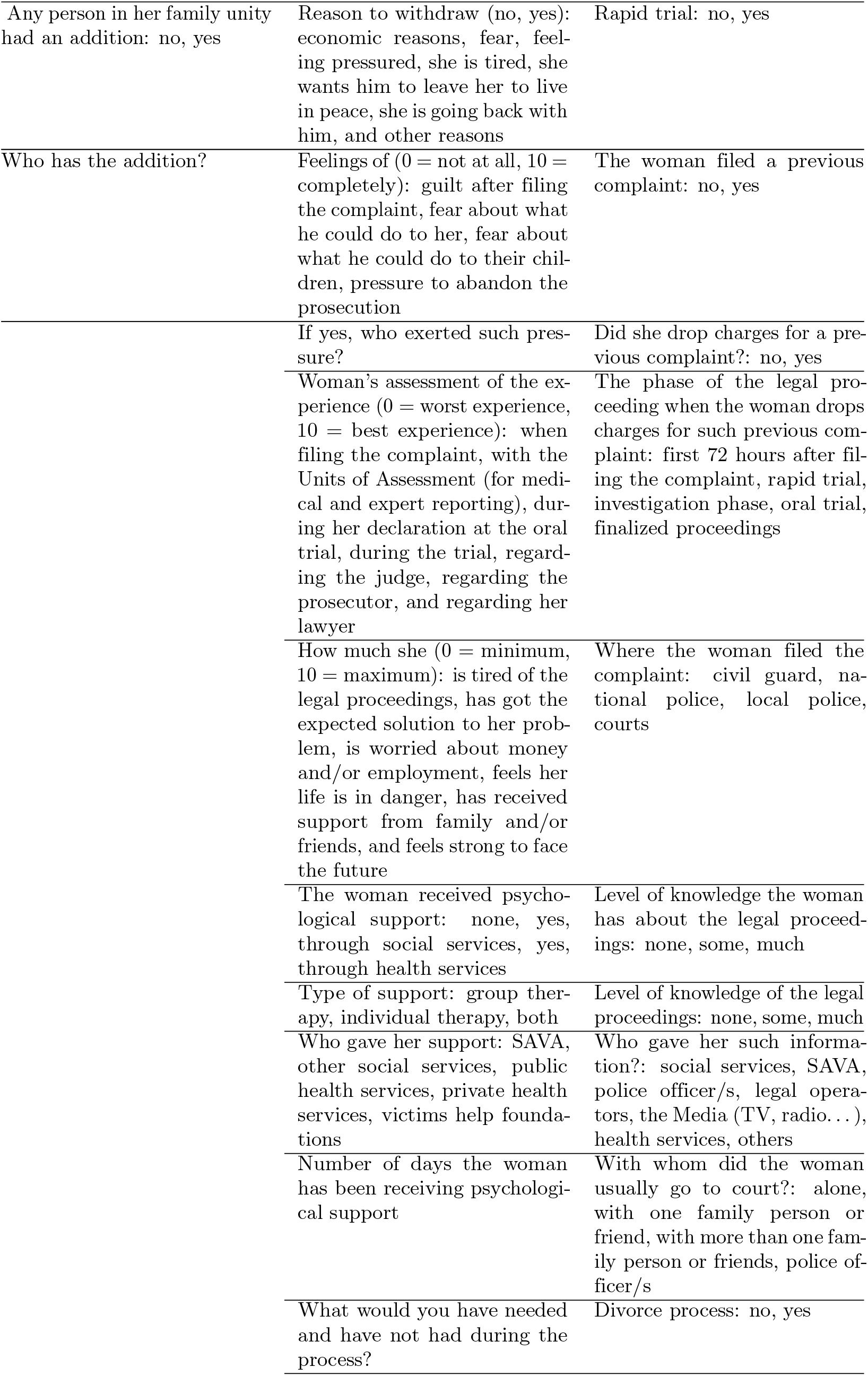
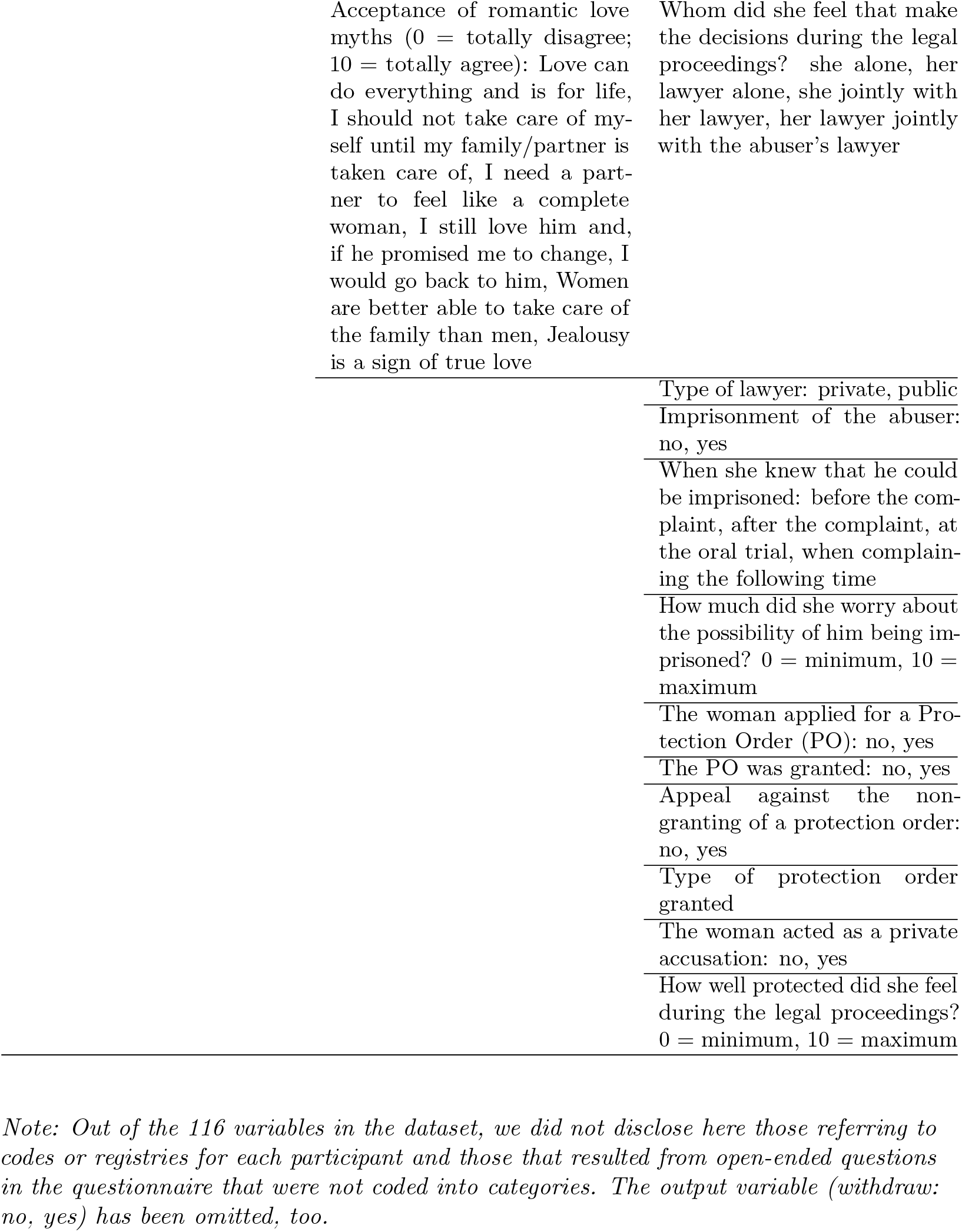

